# Infection Control Program Among the Ysleta del Sur Pueblo Prevents Hospitalizations and Deaths During the Covid-19 Pandemic

**DOI:** 10.1101/2023.12.05.23299531

**Authors:** Cameron M. Torres, Victoria Aparicio, Gabriela Morales, Ascension Mena, Charles T. Spencer

## Abstract

In response to the SARS-CoV-2 pandemic, the United States declared a state of emergency and implemented large scale shutdowns and public health initiatives to prevent overwhelming public resources. The success of these prevention methods remains unresolved as restrictions and implementation varied greatly from national, state, and local levels. Despite national and local regulations, individual adherence to preventative guidelines presented an additional layer of variability. Cases of Covid-19 continued to rise and fall over a two-year period on a national level despite masking recommendations, ease of testing, and availability of vaccines. The Ysleta del Sur Pueblo is a Native American tribal community and sovereign nation located in El Paso, Texas. Speaking Rock Entertainment Center is a major business operated by the tribe, employing a large number of tribal and non-tribal members from the El Paso community. Following nationwide re-openings of non-essential businesses, Speaking Rock implemented an infection control program with strict adherence to recommendations provided by the Center for Disease Control and Prevention (CDC). Herein, we examine the efficacy of these measures and report on the success of the program resulting in zero employee hospitalizations or deaths in a region heavily impacted by severe Covid-19 during the height of the pandemic.

## 1.1 Emergence of SARS-CoV-2

SARS-CoV-2 belongs to the *Coronaviridae* family of viruses sharing 79% of its genome identity with SARS-CoV and 50% sequence identity with Middle East respiratory syndrome coronavirus (MERS-CoV), both highly virulent zoonotic coronaviruses implicated in fatal respiratory disease in humans. SARS-CoV-2 is spread from human to human via the inhalation of respiratory droplets and aerosols. Entry into susceptible host cells is mediated by the binding of viral spike (S) protein with angiotensin-converting enzyme 2 (ACE2) receptors. Following this interaction, the S protein is cleaved activating the S2 subunit which fuses the lipid bilayers of the virus and the host cell facilitating deposition of the viral RNA genome into the cell. The virus replicates in the upper respiratory tract initially by infecting epithelial cells in the nasal cavity. Migration deeper into the airways and lungs triggers a strong immune response and hyper-cytokinemia syndrome referred to as a cytokine storm. This hyper-inflammation causes acute respiratory distress syndrome (ARDS) and is typically the cause of death in fatal cases of Covid-19.

Within a few months of gaining the attention of public health officials, the virus had spread across the globe with over 118,000 positive cases reported in 114 countries resulting in 4,291 deaths by March of 2020. The staggering scale of the outbreak and highly transmissible nature of the virus led to the World Health Organization (WHO) declaring the crisis a pandemic on March 11, 2020. Days after this announcement, individual states began implementing mandatory shutdowns of public places, schools, and non-essential businesses. During this time, the Center for Disease Control and Prevention (CDC) issued a number of preventative measures and guidelines including masking and social distancing recommendations. By early April 2020, the United States led the world in both confirmed cases (500,000) and deaths (18,600) attributed to Covid-19.

## 1.2 Community Impact of Covid-19

In west Texas, the city of El Paso and the surrounding communities within El Paso County (1,015 sq mi) were severely affected by the virus throughout the pandemic. The population of 865,657 predominantly reside within 25 zip codes, 79821-79938. At the time of publication, El Paso County had seen 19,949 hospitalizations and 3,691 deaths as a result of COVID-19, with >50% of hospitalizations and deaths from those residing within 7 zip codes: **79907**, 79912, 79925, **79927**, 79928, 79936, 79938 (Figure 1A,B). These regions constitute the heaviest burden of SAR-CoV-2 in the El Paso community.

**Figure 1.**
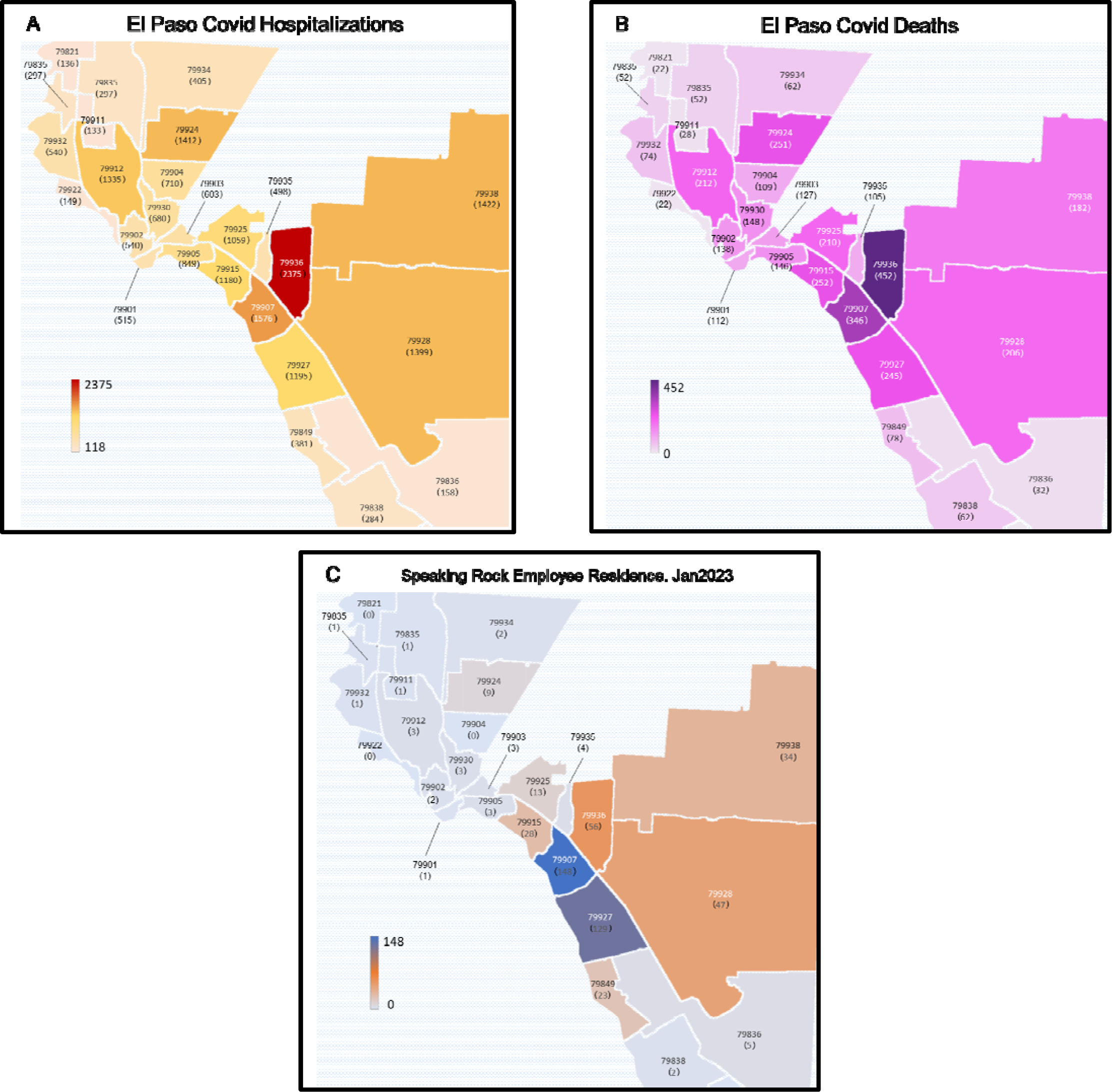
Geographic distribution of severe Covid and residence of Speaking Rock employees within El Paso County. Regions severely impacted by Covid-19 resulting in high incidence of hospitalizations (A) coincide with areas of high Covid-related deaths (B). The majority, >80%, of Speaking Rock employees reside within these hotspots (C).

The Ysleta del Sur Pueblo is a federally recognized Native American tribe (∼4,696 members) of the Tigua peoples situated geographically within the **79907** and **79927** zip codes in El Paso, Texas. The Speaking Rock Entertainment Center (SREC) is a major tribal entity that provides employment opportunities for members of the pueblo as well as the wider El Paso community. At the time of this publication, Speaking Rock employed ∼520 persons in a variety of positions. Of these, 82.22% of employees resided in areas with the highest rates of Covid-19 hospitalizations and deaths in the general El Paso area. As the Ysleta del Sur Pueblo are a sovereign governing body located within the El Paso county area, this provided a unique opportunity to directly compare two sets of epidemiological control measures for an interacting group of individuals in an isolated geographic space.

Following national shutdowns of non-essential businesses, Speaking Rock Entertainment Center closed its doors mid-March 2020. It would re-open in May 2020, with reduced staffing and a rigid infection control program in place to exceed guidelines set by the Center for Disease Control and Prevention (CDC). The epidemiology of SARS-CoV-2 within this small community of employees would be significantly affected by the control measures taken within this program as evidenced by the lack of severe morbidity and mortality among SREC employees when evaluated against the representative Ysleta del Sur Pueblo and El Paso County communities.

## 2.1 Infection Control Program

As a sovereign nation, the Ysleta del Sur Pueblo and Speaking Rock Entertainment Center had authority to enact and enforce a public health program independent of that in the surrounding city, county and state governances. Speaking Rock Entertainment Center has an onsite Wellness Response Department with a medical doctor and nurses available to the employees and their families. At the onset of the Covid-19 pandemic, the Wellness Response Department was charged with designing and instituting infection control protocols with the aim of stemming the spread of the virus. The program enacted by the Wellness Response Department at Speaking Rock Entertainment Center consisted of sterilization equipment, facility upgrades and medical surveillance to reduce the incidence and spread of SARS-CoV-2 amongst employees, families, visitors, and the community as a whole. As the facility was reopened in May 2020, commercial air purifiers and surface purifiers were installed along with sterilization tunnels at all entrances. The entire facility was sprayed with antimicrobial solution three times a day. Partitions between individual stations separated patrons and mandatory mask mandates were implemented for all guests and employees. Onsite PCR testing was made available to employees and their families by mid-May 2020 and the testing center was open 16 hours a day, 7 days a week. This step negated the need to test at a city testing center. If any employee did test positive at an external testing center, they would be re-tested at the center to confirm the results of the previous test. Wait times for the results of PCR tests were on par with the city of El Paso, initially 2-3 days, but then was significantly lengthened, 10-12 days, as testing demand increased and ultimately decreased to 7 days turn around. Employees who tested positive were quarantined for 14 days early in pandemic response. This was later reduced to 7 days following recommendations by the CDC. As test result wait times increased due to increased testing numbers, the center transitioned to rapid antigen testing in December 2020. If an employee or family member tested positive with a rapid test, they would be tested by PCR to confirm the positive result. Correlation between positive rapid tests and PCR confirmation was 100%.

## 2.2 Employee Health Management

While the entertainment center is located within the Ysleta del Sur Pueblo, it does not employ only tribal members with 47.04% of employees outside of the tribe (Table 1). Collectively, the majority (82.22%) of its employees reside in the nearby 79907 and 79927 zip codes (Figure 1C). In addition, it serves the entire El Paso metropolitan area including all of El Paso County, southern New Mexico and Ciudad Juarez, Mexico providing a high chance of exposure to infectious diseases to employees. This was of particular concern during the SARS-CoV-2 pandemic 2020-2021.

**Table 1.** Demographics and underlying health conditions of current employees of the Speaking Rock Entertainment Center. Tribal members make up a little over half of employees. Medical data shown represents the incidence of underlying health conditions associated with severe SARS complications.

|  |  |  |  |
| --- | --- | --- | --- |
| Table 1. |  |  |  |
| Demographics of Speaking Rock employees |  |  |  |
|  | Male | Female | Total |
| Tribal members | 27.30% | 25.60% | 52.90% |
| Non-tribal members | 24.80% | 22.30% | 47.10% |
| Comorbidities among Covid positive employees and El Paso Covid-related deaths |  |  |  |
|  | SREC total (523) | SREC Covid + | EP Covid deaths |
| Hypertension | 15.47% | 16.90% | 10.00% |
| Diabetic/pre-diabetic | 10.69% | 14.70% | 11.00% |
| Chronic Lung disease | 7.07% | 8.77% | 6.00% |
| Cardiovascular disease | 5.16% | 4.70% | 6.00% |

The medical staff of the Wellness Response Department tend to the health of employees at SREC as well as maintaining medical records and documenting incidence of infection. Employee health data including infection rates, household exposures, vaccination status, and health conditions were collected throughout the pandemic. The frequency of underlying medical conditions associated with increased SARS-related mortality and morbidity were similar amongst the entire staff, those that tested Covid-positive, and Covid-related deaths in the general El Paso area, respectively (Table I): hypertension (15.5, 16.9, 10%), chronic lung disease (7.07, 8.8, 6%), diabetic/pre-diabetic (10.7, 14.7, 11%), and cardiovascular disease (5.2, 4.7, 6%).

A critical aspect of the Covid mitigation program was the availability of testing services to both employees and their family members as well as the tracking and monitoring of subsequent exposures to Covid-positive relatives outside of Speaking Rock. Incidence of disease among employees indicated that younger males (aged 30-39 and 20-29) had the highest incidence of disease which declined rapidly in older males (Figure 2A). Interestingly, among female employees the disease incidence was similar across a wide age range (20-59).

**Figure 2.**
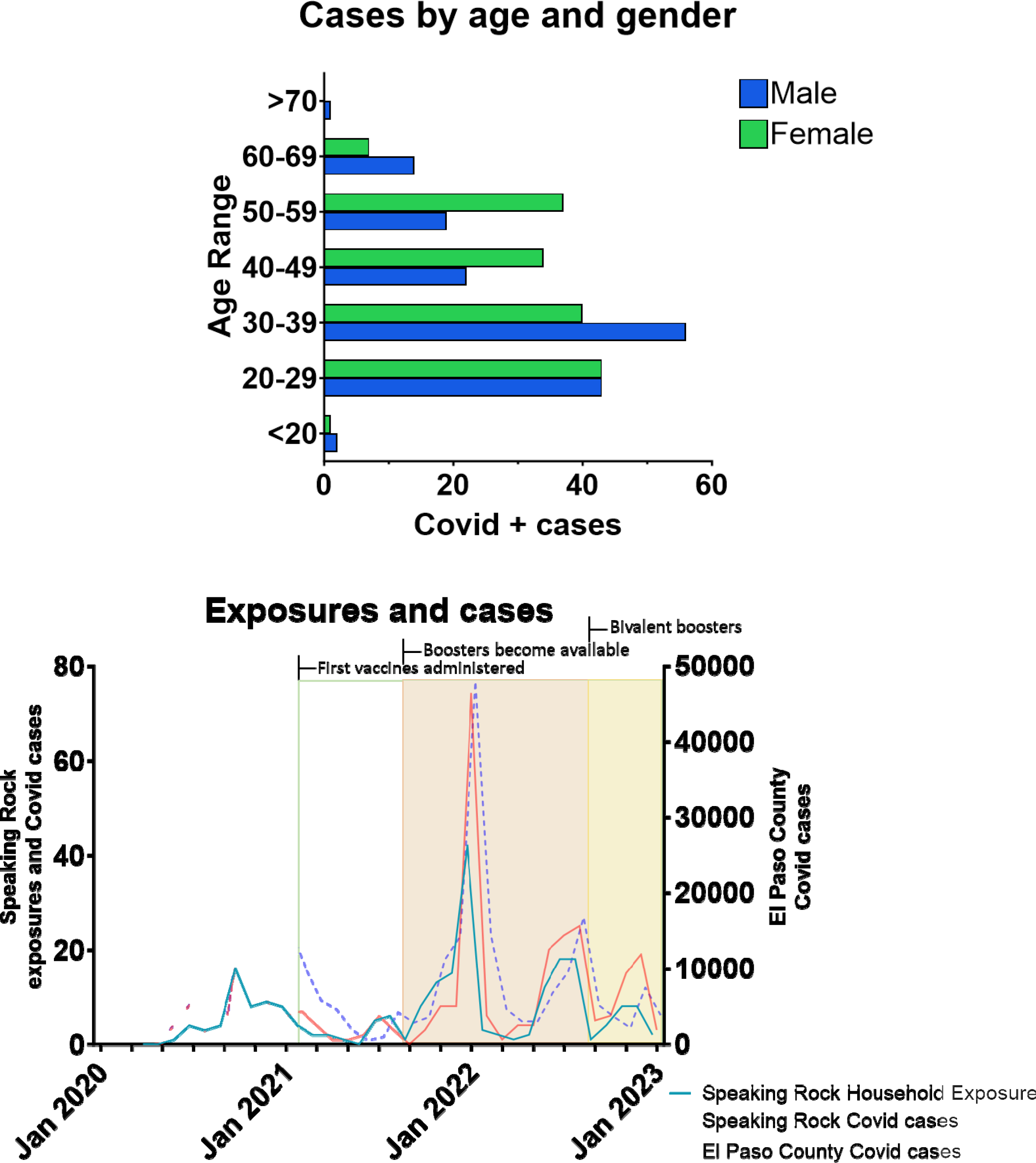
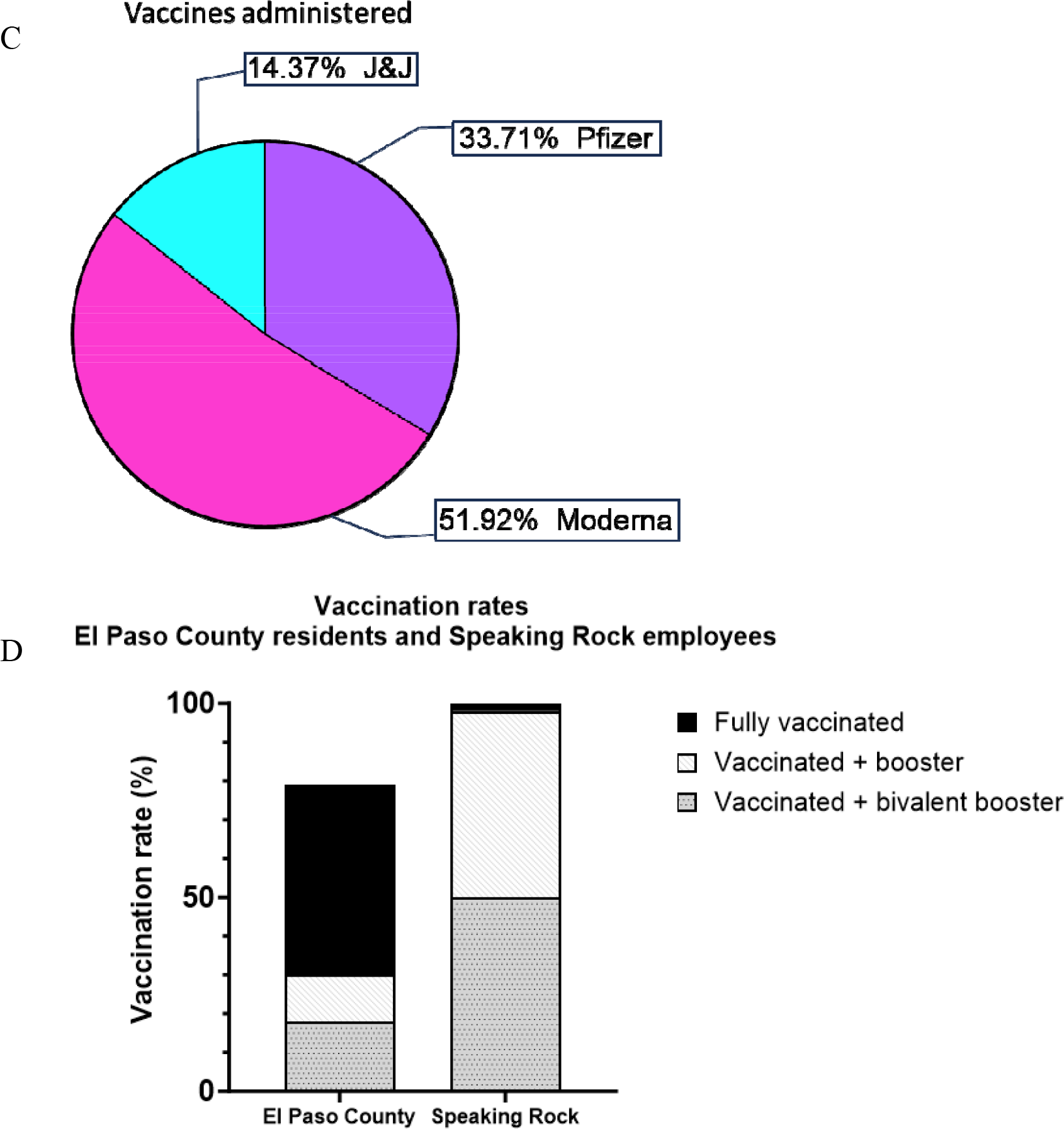
Infection control program designed to detect, track, and prevent the spread of Covid-19. Panels A & B represent testing and tracking data. Panels C & D show data on the vaccines received. The majority of positive cases (A) is shown to be among employees 39 and under, with males constituting the highest rates of positive tests. Exposure rates outside of the workplace (B) coincide with incidence of employees testing positive and suggest transmission to not be associated with employee-to-employee contact. Almost 90% of vaccines received were mRNA vaccines (C), and nearly half the staff have received a Bivalent booster (D).

Covid-positivity of both SREC employees and household contacts generally mirrored the greater El Paso area community transmission rates (Figure 2B) throughout the pandemic with the notable exception in December 2020 to February 2021 where there were disproportionately lower numbers of Covid positive cases amongst SREC employees and their families. This demonstrates the effective control by the prevention strategies of the Speaking Rock Wellness and Response Department early in the pandemic prior to introduction of vaccination. Indeed, detailed analysis of employee and familiar contact cases revealed that Covid positivity was first observed in household contacts. This was then followed by a concomitant rise in employee cases. These data suggest that the majority of Covid cases in employees of Speaking Rock occurred due to community transmission to household members who then transmitted the infection to direct employees with limited transmission within the Entertainment Center, again representing the success of their prevention and control activities.

As the business of a sovereign nation, SREC ensured that all medically competent employees be vaccinated against SARS-CoV-2 as effective vaccine candidates became available in January 2021 with 98.5% coverage (Figure 3). Employees of Speaking Rock were subsequently administered a variety of effective vaccines of which 85% were mRNA-based vaccines requiring the bivalent booster (Figure 2C). By June 2021, all staff were fully vaccinated and all new hires would receive their first doses upon hiring.

The initial booster was also made mandatory regardless of which vaccine was received. Subsequent boosters were not mandated but were strongly encouraged and made readily available. Primary and secondary doses as well as boosters were administered on an employee’s last day of work in a given week to ensure they would be home should any side effects occur. Once Bivalent boosters were developed, which provide a broad spectrum of protection across a range of Covid variants, they were made available to staff. At present time, nearly 50% (well above national rates) of employees have received the Bivalent booster. With the CDC trending towards recommending yearly Bivalent vaccines, it is likely that the health department at SREC will make it a yearly requirement along with the influenza vaccine. Vaccination and booster rates at speaking rock exceed those in the surrounding area of El Paso County (Figure 2D) with roughly 98% of employees being fully vaccinated and having received at least one booster shot. While El Paso county reports 31% fully vaccinated and boosted, and rates of bivalent boosters at roughly 18% nearly a year after their introduction.

## 3.1 Impact of Disease Control Measures

The public health burden of Covid-19 has been quite severe on the El Paso community with 38% of the population testing positive for SARS-CoV-2 infection. Of those, nearly 20,000 cases resulted in hospitalizations with 3,700 deaths (Figure 3A). Officials report just under 80% of the population have been fully vaccinated since the beginning of the pandemic. By contrast, 98.5% of SREC employees have been fully vaccinated with 100% having received at least the first vaccination. Despite 320 Covid positive cases among the employees of Speaking Rock Entertainment Center, none of them resulted in severe disease requiring hospitalization and there were no deaths (Figure 3B). This comparison is significant in that, death and hospitalization rates in the surrounding community are 1 in 250, and 1 in 40 respectively. Taken together, this demonstrates the effectiveness of intense prevention and control program implemented by the Wellness and Response Department of Speaking Rock Entertainment Center.

## 3.2 Discussion

The impact of SARS-CoV-2 has been felt across the globe over the past 3 years. Different communities have been disparately affected by the pandemic and the profound economic burden. Despite the advances in vaccine development and medical technology, vaccine hesitancy, social disparities, and resistance to medical science have exacerbated the toll this virus has had on human health. Apparent from the SARS-CoV-2 pandemic was a lack of cooperation by the general populace for the greater public health.

In this report, we highlight a disease mitigation program that has effectively prevented morbidity and mortality in a region hard hit by the Covid pandemic. This program exceeded recommendations established by the Center for Disease Control and Prevention and, through that program, effectively prevented spread of disease amongst its population. Indeed, compared with the surround are of El Paso County, Speaking Rock Entertainment Center saw no hospitalizations or deaths associated with the pandemic following reopening.

Mitigation procedures were immediately implemented upon reopening of nonessential business and included mandatory masking, sterilization tunnels, surface and air sanitation. Once available, the program expanded to include mandatory vaccination. The success of this program underscores the importance of intensive infectious disease control in public health programs and policies and highlights the need for cooperation among the populace. Indeed, the sovereign nation of the Ysleta del Sur Pueblo allowed them to implement their own mitigation strategy whereas the remainder of El Paso County followed regulations put in place by the State of Texas. This unique geographical arrangement allowed a direct comparison of these mitigation strategies for an intermingling population in the same area. The results from this mitigation strategy encourages widespread access to healthcare and infection monitoring in reducing the community burden of severe disease.

## Data Availability

All data produced in the present study are available upon reasonable request to the authors

## Supplemental Figures

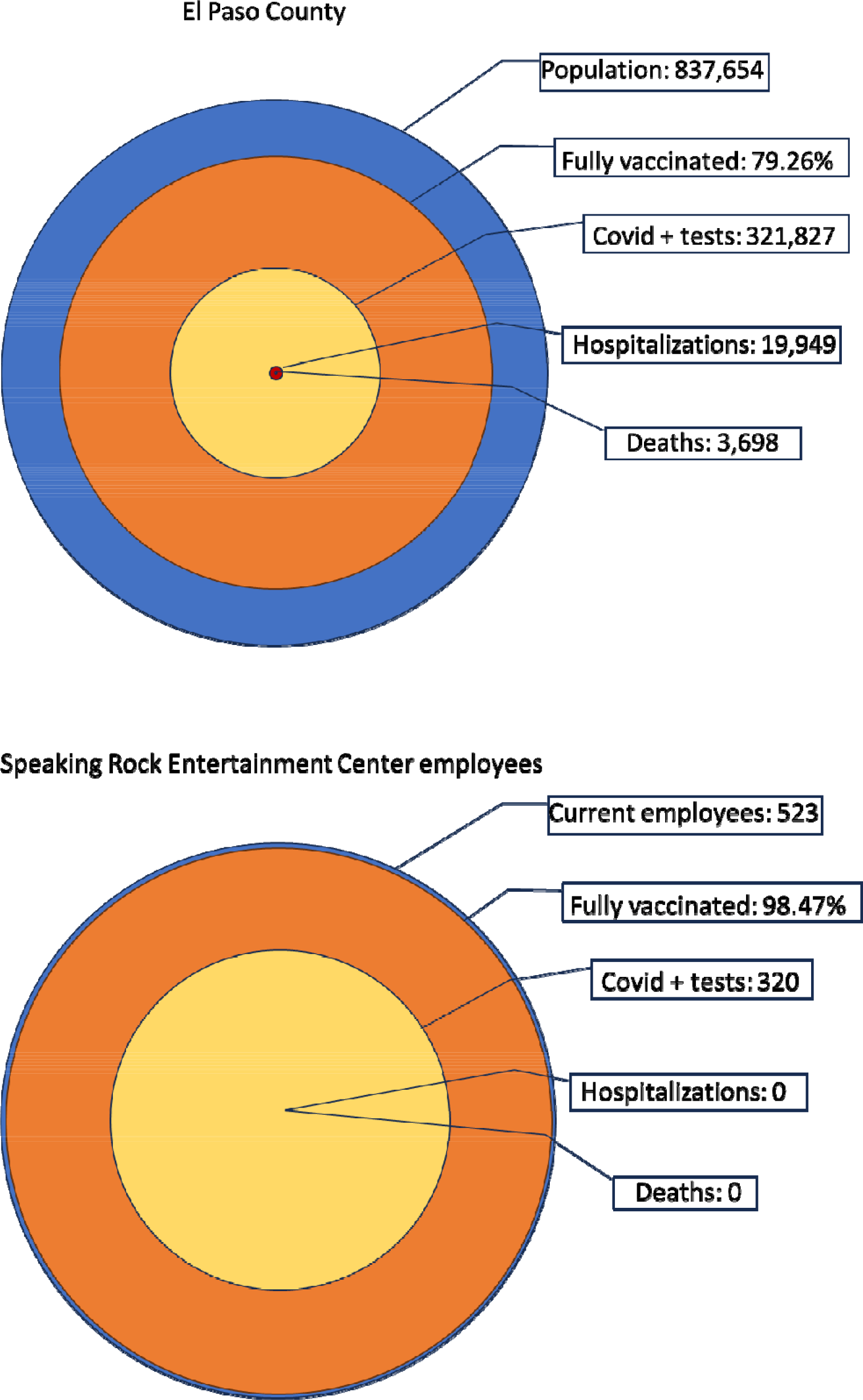

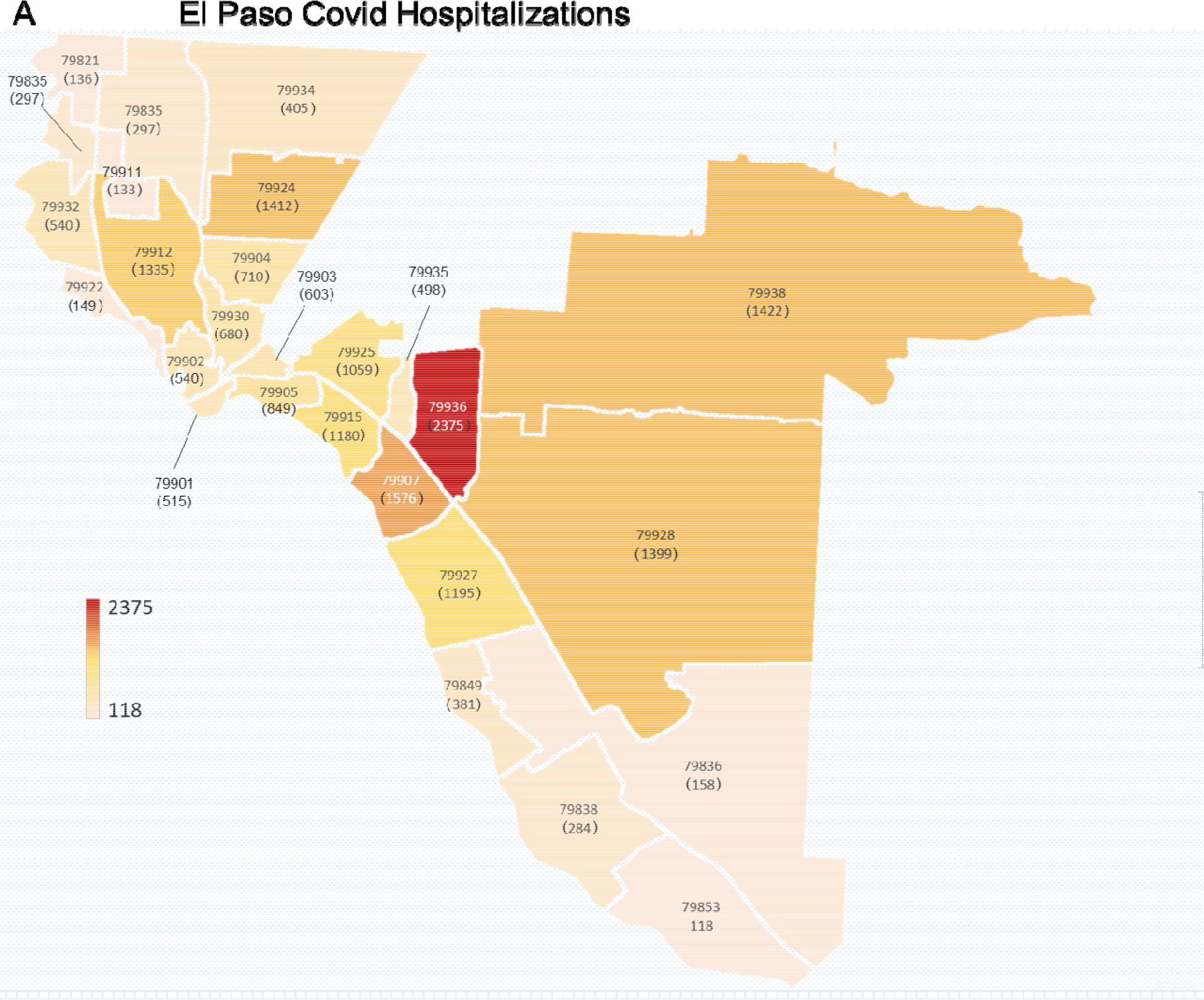
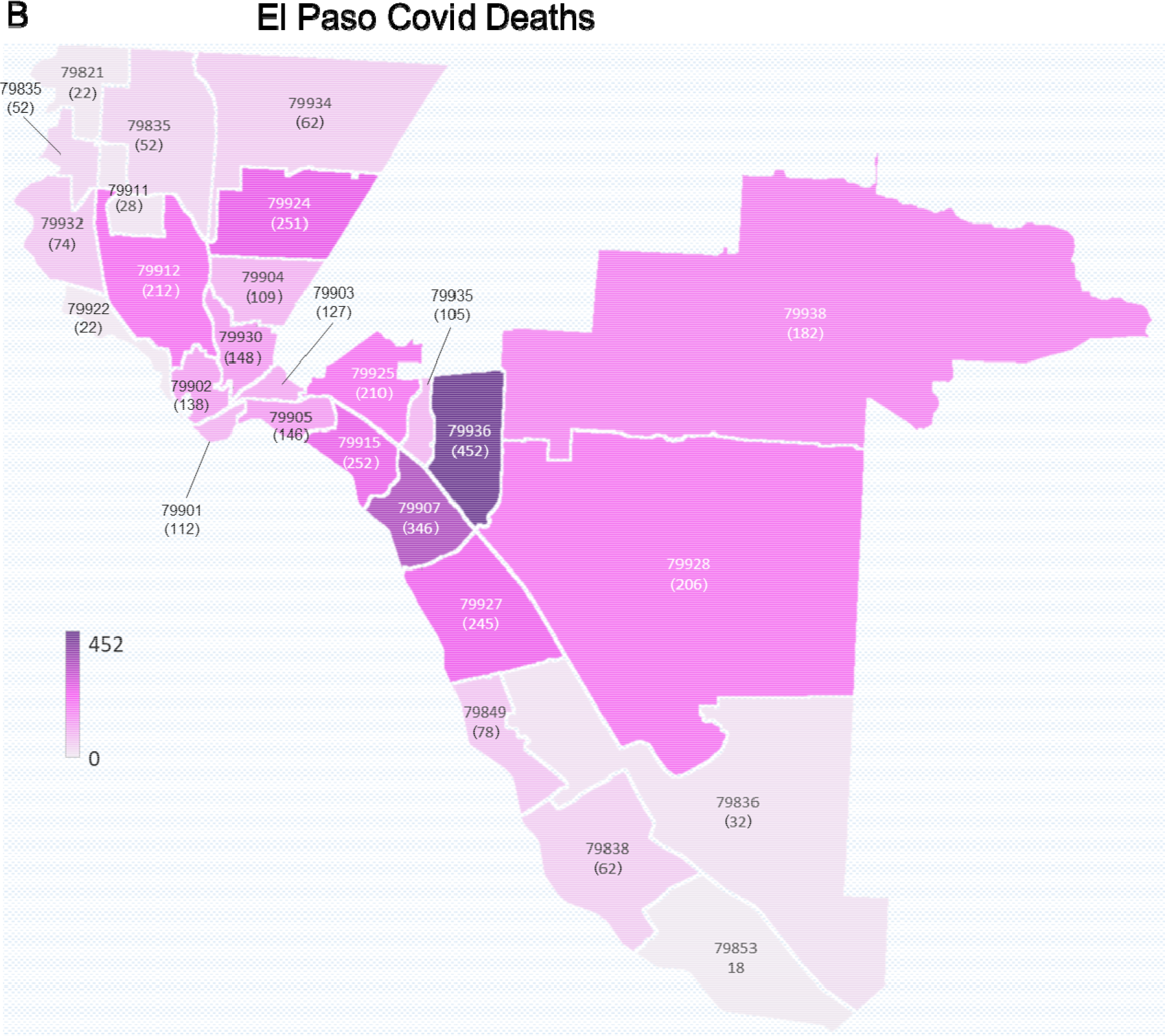
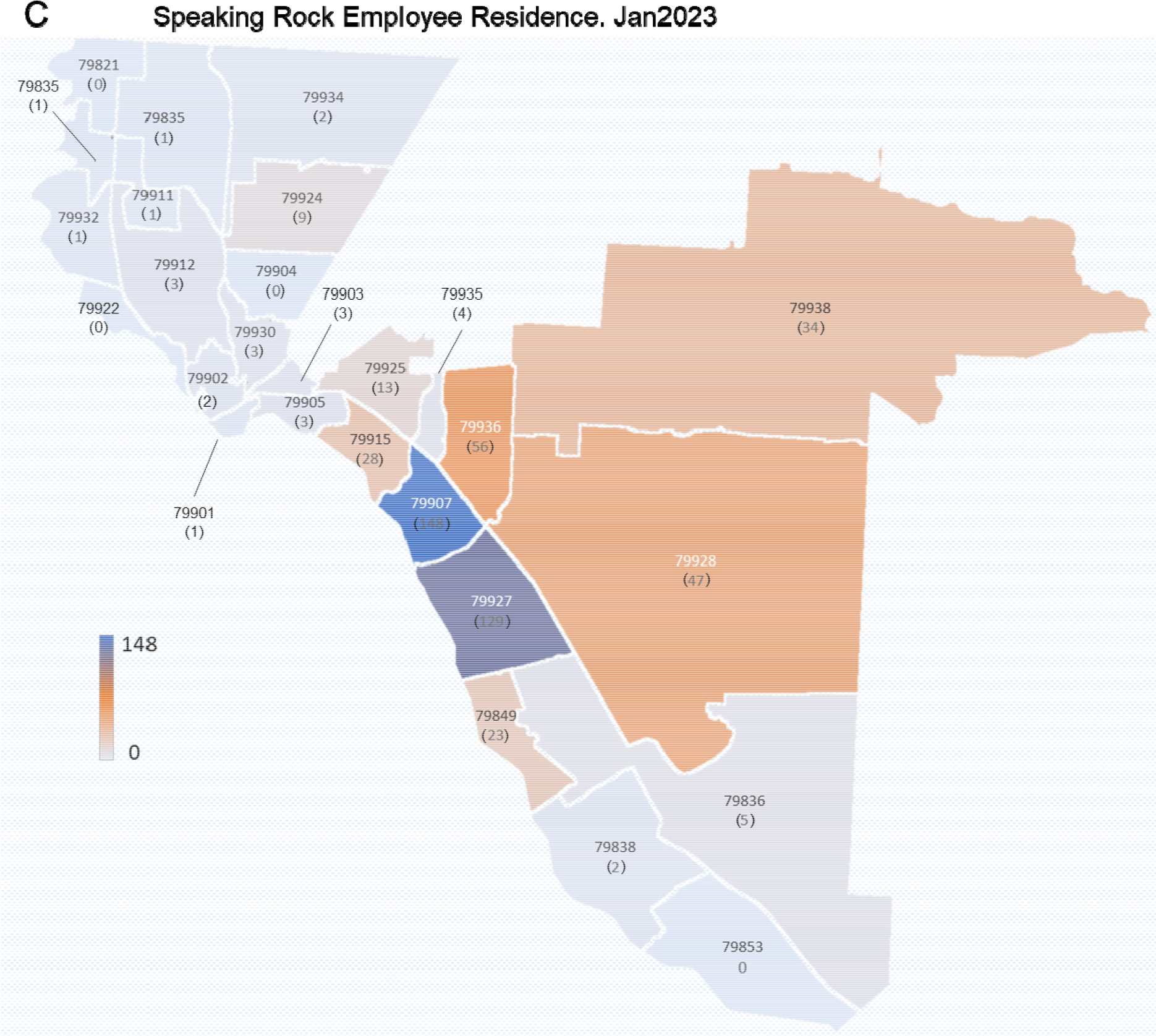

## Notes

**Conflict of Interest Statement:** This report presents data collected by the Wellness Response Department of Speaking Rock Entertainment Group. The data was collected by VA, GM, and AM as employees of Speaking Rock Entertainment Group. The data was analyzed by CMT with one semester funding from Speaking Rock Entertainment Group. Publication costs were partially covered by Speaking Rock Entertainment Group. Dr. Spencer received no direct or indirect funding from Speaking Rock Entertainment and oversaw the validity of this report.

### Competing Interest Statement

This report presents data collected by the Wellness Response Department of Speaking Rock Entertainment Group. The data was collected by VA, GM, and AM as employees of Speaking Rock Entertainment Group. The data was analyzed by CMT with one semester funding from Speaking Rock Entertainment Group. Publication costs were partially covered by Speaking Rock Entertainment Group. Dr. Spencer received no direct or indirect funding from Speaking Rock Entertainment and oversaw the validity of this report.

### Funding Statement

This study was funded by Speaking Rock Entertainment Center.

### Author Declarations

This work was reviewed and approved by the Institutional Review Board at the University of Texas at El Paso.

